# Prevalence of pesticides in European water and relationship for public health: A protocol for systematic review and meta-analysis

**DOI:** 10.1101/2023.02.15.23285961

**Authors:** Manuel Serrano Valera, Nuria Vela, Grasiela Piuvezam, Francisco Mateo-Ramírez, Isaac Davidson Santiago Fernandes Pimenta, Isabel Martínez-Alcalá

## Abstract

**Background:** There is currently a growing interest in the so-called emerging pollutants, such as pesticides, pharmaceuticals, personal hygiene care products, drugs, etc., whose presence in natural ecosystems is not necessarily recent, but the development in latest years of new and more sensitive methods of analysis have allowed their detection. They can be present in the natural environment, in food, and in many products of everyday origin, which suggests that human exposure is massive and universal. These contaminants correspond, in most cases, to unregulated substances, which may be candidates for future regulations, depending on research on their potential health effects and monitoring data regarding their prevalence. Therefore, the study of this type of substances is becoming one of the priority lines of research of the main agencies dedicated to the protection of public and environmental health, such as the World Health Organization (WHO), the United States Environmental Protection Agency (USEPA) or the European Union (EU). In this sense, it is of vital importance to know the nature and quantity of this type of contaminants, to establish preventive mechanisms that minimize their presence in the environment, including, of course, aquatic systems, with special requirements for water human consumption.

**Methods:** This systematic review and meta-analyzes protocol is conducted using the Preferred Reporting Items for Systematic Reviews and Meta-Analyzes (PRISMA) statement guidelines and the Cochrane Handbook of Systematic Reviews of Interventions. The papers should include monitoring studies carried out in pesticides polluted waters in Europe. Prevalence studies of emerging contaminants (pesticides) in water resources (watersheds, aquifers, river, marine and springs), waste waters (influent and effluent) and drinking water should be included. Therefore, the comprehensive search strategy will be conducted in the following databases: PubMed, Scopus, Web of Science, EMBASE, ScienceDirect. Two independent reviewers will conduct all study selection procedures, data extraction, and methodological evaluation, and disagreements will be referred to a third reviewer. RevMan 5.3 software will be used to gather data and perform the meta-analysis if possible.

**Discussion:** This systematic review should provide evidence on the presence of pesticides in European waters in order to establish preventive mechanisms that minimize their presence in the environment.

**Dissemination and ethics:** The findings of this scoping review will be disseminated in print, at conferences, or via peer-reviewed journals. Ethical approval is not necessary as this paper does not involve patient data.

**Systematic review registration:** PROSPERO CRD42022348332.

## 1.- Introduction

Water scarcity now affects all continents and constitutes one of the major challenges of the 21st century. Over the last years, water use and consumption grew at twice the rate of population growth and, although there is enough drinking water on the planet, its distribution is irregular, it is wasted, contaminated in many cases, and is often unsustainably managed [1]. The problem of supply is exacerbated by the fact that a large proportion of surface and groundwater is polluted by the discharge of waste generated by human activity. The United Nations estimates that every day 2 million tons of wastes are disposed of through watercourses. This reduces the availability of drinking water and makes it necessary to resort to treatment methods. Mankind’s control over these waters is now global and human beings now plays an important role in the water cycle. Although the measures taken in the last century to prevent environmental pollution have drastically reduced the presence of some pollutants, such as the so-called “persistent organic compounds”, the number of known potentially dangerous substances that can affect the environment, and therefore man, is still very large. Research attention has been focused on the so-called “emerging pollutants” (ECs). ECs are natural or synthetic chemical substances of various etiologies whose presence in natural ecosystems is not necessarily new, although the development in recent years of new and more sensitive methods of analysis has made it possible to warn of their presence, so that concern about the possible consequences for human health has increased [2]. ECs occur in many kinds of daily chemicals including pesticides (used to protect crops from insects, weeds, fungi and other pests), disinfection by-products (swimming and drinking water purification such as chlorinated by-products), plasticizers (used to food packaging such as phthalates and bisphenols), human and veterinary pharmaceuticals (analgesics, antimicrobials, antidiabetics, antibiotics, antihistamine, or antiepileptics), manufactured nanomaterial (used in information technologies such as silicon dioxide and carbon nanotubes), and finally, preservatives and ultraviolet filters (used to personal care products such as parabens or benzophenones) [3].

The high risk lies in the fact that environmental and human toxicological information for most of the ECs have not been studied yet, therefore it is almost impossible to predict the health effects on living creatures [4]. In addition, the suspected effects could even be intensified on vulnerable groups such as infants and pregnant women as the damage in the very early development stages is significant [5].

Human and his environment are in contact with all these compounds, both from the moment of their manufacture and through the processes of distribution, use and final degradation. Also of concern is the expected increase in their number and concentration, due to the greater reuse of urban, agricultural and industrial wastewater, given the greater need for the availability of drinking water, which means that the study of this type of substances is becoming one of the priority lines of research of the main bodies dedicated to the protection of public and environmental health, such as the WHO, the USEPA and the EU. It should be noted that most ECs are not regulated, so they may be candidates for future regulation, depending on research on their potential health effects and monitoring data regarding their incidence.

On the other hand, the requirement to produce enough food to supply the world’s population has had an impact on agricultural practices around the world. In many countries, this pressure has led to an expansion of irrigation and an increasing use of fertilizers and pesticides to achieve and maintain higher yields. It should be noted that agriculture is the largest user of freshwater resources, using a global average of 70% of all surface water supplies. Agriculture is both a cause and a victim of pollution of water resources. It is a cause, through the discharge of pollutants and sediments into surface and/or groundwater and through the salinization and waterlogging of irrigated land. It is victimized using wastewater and contaminated surface and groundwater, which in turn affect crops and transmit diseases to consumers and agricultural workers. The EU establishes a framework for the prevention and control of water pollution that includes both measures to assess the chemical status of water and measures to reduce the presence of pollutants. As a matter of priority, as set out in Directive 2013/39/EU of 12 August 2013 amending Directives 2000/60/EC and 2008/105/EC as regards priority substances in the field of water policy [6], the causes of pollution must be identified and pollutant emissions must be treated at the source itself, in the most economically and environmentally effective way. Some strategies were agreed in the Millennium Development Goals through the Sustainable Development Goals (SDGs), specifically the goal 6 “Ensure availability and sustainable management of water and sanitation for all” [7].

This fact is evident in the results of numerous worldwide monitoring studies carried out on wastewater in which a wide range of ECs have been found at different concentration levels, among which pesticides stand out for their high incidence [8-10].

This group of emerging contaminants is of particular concern to agencies such as WHO and European Food Safety Authority (EFSA). Pesticides are very important for food production, as they maintain and/or increase crop yields, which is especially important in countries suffering from food shortages. However, most of them are highly persistent in the environment, and can be highly toxic to humans. There is scientific evidence that exposure to pesticides can lead to various health problems such as immunosuppression, hormone disruption, reduced intelligence, reproductive distortion and cancer. In humans, the effects of pesticide exposure can be classified into acute health problems and chronic health problems. Chronic problems include references to neurological effects such as the onset of Parkinson’s disease, reduced attention span, memory impairment, reproductive problems, impaired child development, birth defects and cancer. Acute effects include reduced vision, headaches, salivation, diarrhea, nausea, vomiting, wheezing, coma and even death. Moderate pesticide poisoning can cause intrinsic asthma, bronchitis and gastroenteritis [9]. These effects will depend on the chemical nature of the compound, the amount and the mode of exposure (ingestion, inhalation or direct skin contact).

The use of these substances in agriculture is regulated in developed countries, but unfortunately there is no such rigorous control in many other countries. Extensive EU legislation regulates the placing on the market and use of plant protection products and their residues in food. EFSA evaluates active substances used in plant protection products by providing scientific advice on toxicological risks. The risk assessment of active substances analyzes whether, when used correctly, these substances are likely to have any direct or indirect harmful effects on human or animal health, for example, through drinking water, food or feed, or on groundwater quality.

The EU and the Member States take regulatory risk management decisions, including the approval of active substances and the establishment of legal limits for pesticide residues in food and feed (maximum residue levels or MRLs).

Despite all this regulation by the competent authorities, the presence of pesticide residues in aquatic ecosystems remains high, so it is of vital importance to know the nature and quantity of this type of contaminants, to establish preventive mechanisms to minimize their presence in the environment, including, of course, aquatic systems, with special requirements for water for human consumption.

## 2. Objective

The main objective of this work is to systematically review, analyze and synthesize the available evidence on the status of pesticides in waters in Europe. For this purpose, quantitative studies reporting the prevalence and/or a concentration of several types of pesticides such as glyphosate, chlorpyrifos, pyrethroid pesticides, neonicotinoid pesticides, and/or fungicides, in samples of different water resources like wastewaters and drinking water will be analyzed.

To our knowledge, this is the first systematic review and meta-analysis that collects and organizes the existing evidence on the pesticide’s contamination of water resources [10], to quantify the environmental health status in the waters from Europe.

## 3. Methods and analysis

### 3.1. Study registration

This systematic review has been registered on PROSPERO (CRD42022348332) and will develop in accordance with the Preferred Reporting Items for Systematic Reviews and Meta-Analyses (PRISMA) statement guidelines [11] and the Cochrane Handbook of Systematic Reviews of Interventions [12].

### 3.2. Study selection criteria

#### 3.2.1. Types of studies

Prevalence studies of emerging contaminants (pesticides) should be included.

#### 3.2.2. Types of participants

Water resources (watersheds, aquifers, river, marine and springs) wastewaters (influents and effluents) and drinking waters contaminated by emerging pollutants should be included. The resources will be focused on water contamination by emerging pollutants in Europe.

#### 3.2.3. Types of interventions

No interventions will be made in this study as the objective is to analyze the prevalence of pesticide contamination in European waters based on previously published monitoring data.

#### 3.2.4. Types of outcomes

The results may include: 1. Improvement in the knowledge on the main sources of pesticide contamination in water at European level; 2. Increase in the information on which compounds have the highest frequency of occurrence; 3. Improved knowledge about the level of contamination by these compounds and the areas with the highest contamination load.

### 3.3. Search strategy

This systematic review will summarize evidence published by primary trials through a comprehensive search in the following databases: PubMed, Scopus, EMBASE, Web of Science and ScienceDirect. The search strategy results from a combination of free text search terms and Medical Subject Headings (MeSH), text words, and keywords. The following keywords will be used: word group 1: pesticides AND word group 2: water resources OR wastewater OR drinking water OR water OR sanitation AND word group 3: monitoring AND prevalence, AND word group 4: Europe. For example, the full search strategy for PubMed will be: (wastewater OR water resources OR drinking water OR sanitation AND pesticides).

The search terms used for the formation of the search equations will be combined with specific filters for each database. There will be limitations of time the articles selected for this review will be publications from 2015 onwards, as the EU Water Framework Directive (2000/60/EC) requires all Member States to protect and improve water quality to achieve good ecological status by 2015. In this respect, the main aim of this work is to know the current level of pollution at the European level after the implementation of this regulation. There will be no limitations in language in the searches performed.

### 3.4. Study selection

Two reviewers will independently select the studies by scanning titles and summaries and reading full texts if it is necessary according to the predefined eligibility criteria. Any disagreements regarding study selection will be solved by consulting a third reviewer.

#### 3.4.1. Inclusion criteria

Selected articles should report on prevalence studies of emerging contaminants (pesticides). The articles selected for this review will be publications from 2015 onwards, as the EU Water Framework Directive (2000/60/EC) requires all Member States to protect and improve water quality to achieve good ecological status by 2015. In this respect, we want to know the current level of pollution at the European level after the implementation of this regulation.

#### 3.4.2. Exclusion criteria

The studies must not be focused on leachates and stormwaters and be focused on prevalence studies outside of Europe, prevalence studies of other organic contaminants emerging, prevalence studies of microbiological agents, experimental studies at laboratory scale, pilot plant conditions in water treatment plants, drinking water treatment, review studies, toxicity studies of disinfection by-products (DBPs), prevalence studies of pathogenic microorganisms, resistance mechanism studies, toxicological risk assessment studies in animals, human exposure risk studies, analytical methodology studies, emerging contaminants degradation pathway studies, no project purpose.

### 3.5. Data extraction and management

#### 3.5.1. Data management

After extracting the papers in the databases, an initial check will be performed for the presence of duplicates, followed by their proper removal, using the Rayyan QCRI tool like a reference manager.

#### 3.5.2. Selection process

After the extraction of the papers in all databases, an initial check will be made to look for duplicates, following their proper removal using the Rayyan QCRI tool. Database records will be independently screened by two reviewers (MSV and FMR), reading titles and abstracts. The selected studies will then be fully read to assess compliance with the eligibility criteria. A manual search will be performed if any relevant studies are found using the defined search strategies. References cited in articles will be further reviewed to locate any additional relevant articles not retrieved within the primary search. All investigators (MSV, IMA, NV, ILG, RAB, IDSFP, GP) will then review the full text of all eligible studies.

Every step of the study selection will be documented in a flow chart, as per PRISMA guidelines [10]. A complete list of all references retrieved and separate lists for the included and the excluded at each step, with the respective reason for exclusion, will be available as well. The selection of the study is summarized in a PRISMA flow diagram (Figure 1).

**Figure 1.**
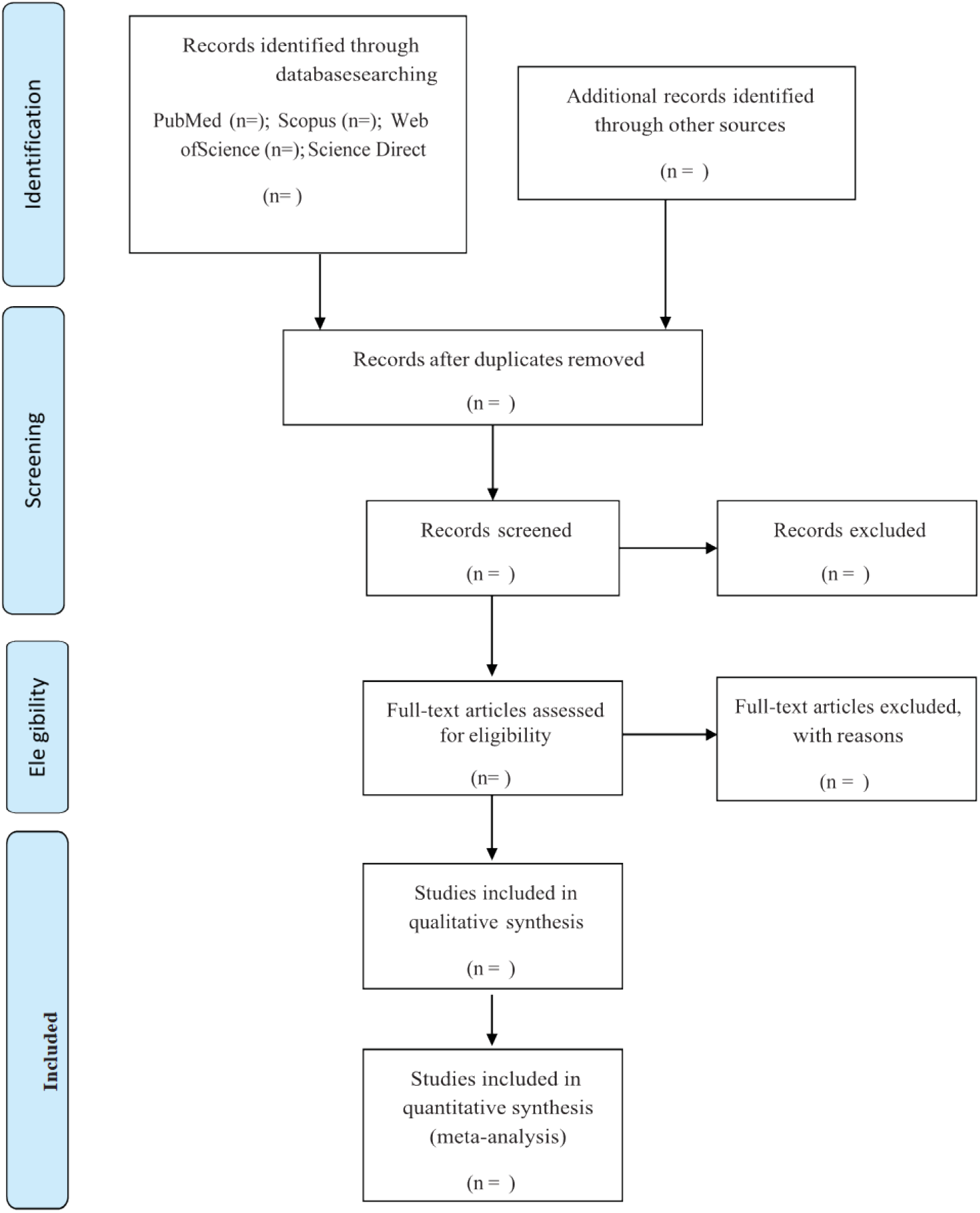
Flow diagram of study selection process.

At this stage, an agreement between the reviewers, regarding the use of the eligibility criteria, will be defined, and the evaluation of the concordance of the investigators will be using the Kappa index. Any disagreements between reviewers in any phase of study selection will be resolved by consulting another reviewer (NV).

#### 3.5.3. Data collection process

A data extraction template in Excel format will be developed and piloted until convergence and agreement among data extractors is reached. The data will be independently extracted by two reviewers (MSV, IDSFP and FMR) from all literature resulted relevant after the screening process. At a minimum, two review authors will independently extract data; A third author (NV) will resolve conflicting extractions, if any.

#### 3.5.4. Data items

We will extract relevant information from each of the selected studies. Figure 2 shows the data that will be extracted from the articles.

**Figure 2.**
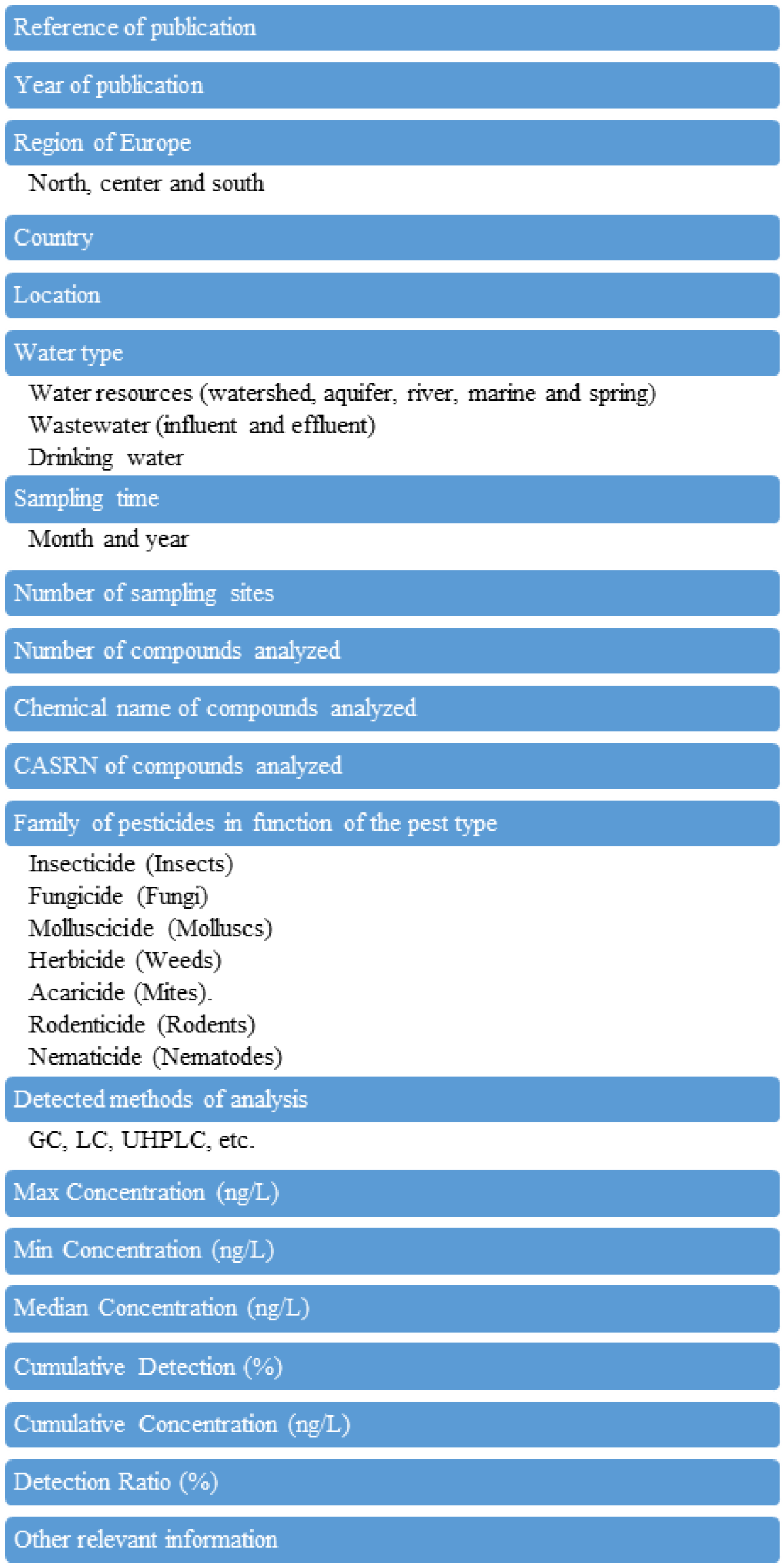
Data extracted from the articles.

If a result is measured at different time points, the mean values and standard deviation will be included.

### 3.6. Dealing with missing data

If the extractions are unclear or incomplete, we will attempt to obtain any missing data by contacting the first or corresponding authors or co-authors of an article via phone, email, or post. If we fail to receive any necessary information, the data will be excluded from our analysis and will be addressed in the discussion section.

### 3.7. Risk of bias assessment

This systematic review will be conducted using the Preferred Reporting Items for Systematic Reviews and Meta-Analyses (PRISMA) guidelines [10]. Two reviewers (MSV, IDSFP and FMR) will independently evaluate the selected studies using the Checklist for Prevalence studies from JBI institute adapted for the included studies in this review [13]. In case of any doubts or discrepancies in both assessments, a third researcher (NV) will be consulted.

### 3.8. Quality of evidence rating

The evaluation of the soundness of the studies will be assessed based on the robustness and sensitivity of the analytical methods used for the determination and analysis of pesticide residues.

### 3.9. Data synthesis

All studies showing concentration data (maximum, minimum and median values) of pesticides in water resources, wastewater and drinking water in Europe will be considered. As far as possible, the following variables will be collected: region of Europe, country, location, type of water, sampling time, number of sampling sites, year of sampling, number of analyzed compounds, chemical name of analyzed compounds, CASRN of analyzed compounds, family of compounds, methods of analysis detected.

Meta-analysis will be performed for the two main outcomes of interest extracted from the studies with the main sources of pesticide contamination in water at European level; with the compounds that have the highest frequency of occurrence and with the level of contamination by these compounds and the areas with the highest contamination load.

Quantitative synthesis of the results will be performed only on studies that analyze drinking water and wastewater; furthermore, meta-analysis will be conducted only on the pesticides that will be found in Europe.

We will produce summary measures and synthesize the evidence quantitatively (i.e., meta-analysis). The type of model to be used for meta-analysis will be chosen based on the evidence. Meta-analysis will be likely performed using random-effects models, since we expect the sets of studies to be heterogeneous in their methods and in the characteristics of the included samples.

Information from all studies that will not be considered suitable for quantitative analysis will be collected in a separate table, so that a qualitative synthesis of their content is available.

Based on the results of the analysis of the proportion of contaminated samples, clusters that define the frequency of the contamination will be created. In particular, the contamination for each pesticide will be defined as:

- Rare: the substance is present in < 1% of the samples.
- Relatively rare: the substance is present in 1–5% of the samples.
- Relatively common: the substance is present in 5–10% of the samples.
- Common: the substance is present in > 10% of the samples

## 4. Discussion

Studies carried out in recent years reveal the presence not only of the well-known ECs but also of numerous variants of these families, in this case the study of pesticides. Pesticides can be found in the water bodies, derived above all, by the runoff from the agricultural field where they have been applied and industrial wastewaters. Especially pesticides more soluble are transported by water molecules during the precipitation event by percolating downward into the soil layers and eventually reaching surface waters and groundwater [14]. Consequently, it produces a degradation of water quality and then reduces the supply of potable water. The presence of pesticides in different environmental compartments, such as groundwater and surface water are a widespread problem. For example, pesticides with high mobility and persistence like organophosphates are detected most frequently in streams and groundwater around the world.

Although the use of pesticides is necessary to get higher agricultural productions, the occurrence of pesticides in the water poses a deleterious effect on human and ecosystem health, where the effect magnitude depends on the pesticide properties such as solubility, half-life, adsorption capacity or biodegradability. The effects produced in human beings can be acute or chronic. Some concrete examples of the effects of the pesticides above human health are reported in literature. The frequent exposure to chlorinated pesticides had a high chance of inducing prostate cancer and allergic or non-allergic asthma in farmers [15,16]. The study of the exposure to the organophosphate pesticides in animals induce genotoxicity effects or induce testosterone and hormone disruption in testis.

In order to protect public and ecosystem health, guideline levels for pesticides in drinking waters have been restricted by governments. For example, there are several guideline values for the presence of pesticides in drinking water issued by the WHO, the United States, Australia, the EU and Japan. In fact, new legislations have been implemented in Europe for the use of regenerated water containing pesticides.

Concerns have increased, as well as the need to detect them and to employ new reduction and elimination techniques. For that reason, this study was established, since to the authors knowledge, although there are many site-specific monitoring studies, there are no studies that conduct a systematic review and meta-analysis of pesticides in Europe.

The study focuses on the systematic review of the status of European Mediterranean waters by meta-analysis of data collected in studies in recent years using the most relevant databases in this field of research. To establish comparative studies between the different regions of the Mediterranean area and the repercussion of these pollutants by families. To offer not only a global vision of the current situation of pharmaceuticals, drugs and personal hygiene products, but also new possibilities to increase the detection and elimination systems of these ECs.

Therefore, it is necessary to resort to what are known as monitoring studies that, through the design and implementation of methods, processes and procedures, guarantee, to a large extent, the follow-up and evaluation of these pesticides, as well as the implementation or execution of observation systems for the ecosystems [17]. In this respect, chemical pollution of natural waters represents a threat to the environment, with effects such as acute or chronic toxicity in aquatic organisms, accumulation of pollutants in the ecosystem and loss of habitats and biodiversity [18, 19]. As a matter of priority, as set out in Directive 2013/39/EU, the causes of pollution must be identified, and pollutant emissions must be dealt with at source in the most economically and environmentally effective way [6]. There are no studies in the literature dealing specifically with this issue and the results of this study will help in the elaboration of public policies for the whole of Europe. The results obtained through this study will contribute to the development of public policies for the whole of Europe and probably for the whole world.

## Data Availability

All relevant data from this study will be made available upon study completion

## Abbreviations

EC: emerging pollutants.
EU: European Union.
PRISMA-P: Preferred Reporting Items for Systematic Reviews and Meta-Analyses Protocols.
USEPA: United States Environmental Protection Agency.
WHO: World Health Organization.

## Author contributions

Manuel Serrano Valera, Isabel Martínez-Alcalá, Nuria Vela, Francisco Mateo Ramírez, Isac Davidson Santiago Fernandes Pimenta and Grasiela Piuvezam.

**Conceptualization:** Isabel Martínez-Alcalá, Nuria Vela, Grasiela Piuvezam.

**Data curation:** Manuel Serrano Valera, Francisco Mateo Ramírez, Isac Davidson Santiago Fernandes Pimenta.

**Formal analysis** Isabel Martínez-Alcalá, Nuria Vela.

**Funding acquisition:** Grasiela Piuvezam.

**Methodology:** Isabel Martínez-Alcalá, Nuria Vela, Grasiela Piuvezam.

**Project administration:** Isabel Martínez-Alcalá, Nuria Vela, Grasiela Piuvezam.

**Resources:** Manuel Serrano Valera, Francisco Mateo Ramírez, Isac Davidson Santiago Fernandes Pimenta

**Supervision:** Isabel Martínez-Alcalá, Nuria Vela, Grasiela Piuvezam.

**Writing – original draft:** Isabel Martínez-Alcalá, Nuria Vela, Grasiela Piuvezam.

**Writing – review & editing:** Manuel Serrano Valera, Isabel Martínez-Alcalá, Nuria Vela, Grasiela Piuvezam.

## Funding

This study was financed in part by the Coordenação de Aperfeiçoamento de Pessoal de Nível Superior - Brasil (CAPES) - Finance Code 001.

## Notes

### Competing Interest Statement

The authors have declared no competing interest.

### Funding Statement

Yes, IDSFP. Finance Code 001. Coordenação de Aperfeiçoamento de Pessoal de Nível Superior. https://www.gov.br/capes/pt-br. Brasil (CAPES). Roled played: Data curation Resources

